# Health Related Quality of Life and Hospitalizations In Chronic Thromboembolic Pulmonary Hypertension Versus Idiopathic Pulmonary Arterial Hypertension: An Analysis from the Pulmonary Hypertension Association (PHAR)

**DOI:** 10.1101/2021.07.21.21260957

**Authors:** Jasleen Minhas, Sai Prasanna Narasimmal, Todd Bull, Teresa De Marco, John Wesley McConnell, Matthew R. Lammi, Thenappan Thenappan, Jeremy Feldman, Jeffrey Sager, David Badesch, John J. Ryan, Daniel Grinnan, Dianne Zwicke, Evelyn Horn, Jean M. Elwing, John E. Moss, Michael Eggert, Oksana A. Shlobin, Robert Frantz, Sonja Bartolome, Stephen C. Mathai, Sula Mazimba, Steven Pugliese, Nadine Al-Naamani, on behalf of the PHAR investigators

## Abstract

Chronic thromboembolic pulmonary hypertension (CTEPH) is a rare, morbid, potentially curable subtype of pulmonary hypertension that negatively impacts health-related quality of life (HRQoL). Little is known about differences in HRQoL and hospitalization between CTEPH patients and idiopathic pulmonary arterial hypertension (IPAH) patients. Using multivariable linear regression and mixed effects models, we examined differences in HRQoL assessed by emPHasis-10 (E10) and SF-12 between CTEPH and IPAH patients in the Pulmonary Hypertension Association Registry, a prospective multicenter cohort of patients newly evaluated at a Pulmonary Hypertension Care Center. Multivariable negative binomial regression models were used to estimate incidence rate ratios (IRR) for hospitalization amongst the two groups. We included 461 IPAH and 169 CTEPH patients. 21% of CTEPH patients underwent pulmonary thrombendarterectomy (PTE) before the end of follow-up. At baseline, patients with CTEPH had significantly worse HRQoL (higher E10 scores) (ß 2.83, SE 1.11, p=0.01); however, differences did not persist over time. CTEPH patients had higher rates of hospitalization (excluding the hospitalization for PTE) compared to IPAH after adjusting for age, sex, body mass index, WHO functional class and six-minute walk distance (IRR 1.66, 95%CI 1.04 – 2.65, p=0.03). CTEPH patients who underwent PTE had improved HRQoL as compared to those that were medically managed, but patients who underwent PTE were younger, had higher cardiac outputs and greater six-minute walk distances. In this large, prospective, multicenter cohort, CTEPH patients had significantly worse baseline HRQoL and higher rates of hospitalizations than those with IPAH. CTEPH patients who underwent PTE had significant improvements in HRQoL.

## Introduction

Chronic thromboembolic pulmonary hypertension (CTEPH) is characterized by remodeling of the pulmonary arteries resulting from acute and recurrent pulmonary emboli with subsequent development of a pulmonary vasculopathy and elevated pulmonary artery pressures resulting in right heart dysfunction. CTEPH leads to functional impairment and negatively impacts health-related quality of life (HRQoL).(1-3) Surgical treatment with pulmonary thromboendarterectomy (PTE) remains the mainstay of treatment for patients eligible for this procedure; however, 20 to 40% of CTEPH patients have inoperable disease or are not surgical candidates for other reasons.(4) One-third of those who undergo PTE still have residual disease.(5, 6) Medical therapies primarily used to treat pulmonary arterial hypertension (PAH) are also used to treat patients with CTEPH; however, the role of medical treatment with or without PTE is not well-defined. At the time of this analysis, riociguat remains the only FDA approved therapy for CTEPH.(7)

In contrast to CTEPH, medical therapies have a well-established role in the treatment of PAH with several FDA-approved drug classes. Despite some improvement with medical therapy, patients with PAH continue to experience significantly worse HRQoL than the general U.S. population. Additionally, patients with PAH have considerably higher rates of health care utilization, including up to four times the cost of matched controls, both before and after their diagnosis.(8-10) Clinical studies often group the diagnoses of IPAH and CTEPH,(11, 12) but, there is limited data assessing patient-reported outcomes in CTEPH and comparing them to other PAH patients.(13) The differences in pathogenesis, treatments and outcomes between these patient populations are important to define in a large generalizable population.

We sought to investigate differences in HRQoL and hospitalizations between patients with CTEPH and idiopathic PAH (IPAH) using data from the Pulmonary Hypertension Association Registry - a large, prospective multicenter cohort of pulmonary hypertension patients in the United States.

## Methods

### Study Sample

The Pulmonary Hypertension Association Registry (PHAR) enrolls patients with PAH or CTEPH who were newly evaluated at one of >50 Pulmonary Hypertension Care Centers in the U.S. The registry began enrollment in 2015 with inclusion and exclusion criteria that have been previously described.(14) For this analysis, we included adult patients (>18 years of age) with CTEPH or IPAH enrolled in the registry between January 2015 (inception of registry) and September 2019. This end date of enrollment was chosen to provide at least 6-months of follow-up before COVID-19 related shutdowns. Patients with both prevalent and incident diagnoses of either CTEPH or IPAH within 6-months of enrollment were included. CTEPH patients who had already undergone PTE before enrollment in the registry (n = 5) were excluded. All subjects were censored at the time of their last follow-up or March 1, 2020 (chosen to avoid the impact of COVID-19 shutdown).

### Clinical Variables

After providing informed consent, demographics, clinical and social history, measurements of exercise performance and hemodynamics via right heart catheterizations were recorded at an initial visit for each patient. Patients were then followed at approximately six-month intervals. At each subsequent visit, patients provided updated demographics, social histories and reported all-cause hospitalizations since their last visit; however, the reason for hospital admission is not recorded in PHAR. Patients also reported symptoms and filled out HRQoL questionnaires described below. In addition, each participating site tracked outcomes, including PTE, lung transplantations, and death.(15) At the time of analysis, the registry had not yet started collecting data on pulmonary balloon angioplasty (BPA).

### Study outcomes

HRQoL was assessed at the initial visit and longitudinally at all follow-up visits using two questionnaires: the medical outcome short form-12 (SF-12), an abbreviated version of the Medical Outcome Study Short Form-36, and the emPHasis-10 (E10), a pulmonary hypertension specific instrument. The SF-12 consists of a physical component score and mental component score. Each component has scores ranging from 0 – 100, with higher scores indicating better HRQoL.(16, 17) The E10 score ranges from 0 – 50 with lower scores indicating better HRQoL.(18)

### Statistical analysis

Baseline patient characteristics were summarized using descriptive statistics: numbers and percentages, mean and standard deviations, and median and interquartile ranges, as appropriate. Multivariable linear regression models were used to assess baseline differences in HRQoL scores in the CTEPH group compared to those with IPAH. These models were adjusted for *a priori* selected potential confounders which included age, sex, body mass index (BMI), baseline six-minute walk distance and their baseline World Health Organization (WHO) functional class. These variables were selected because of their known impact on HRQoL and as surrogates for disease severity. We conducted a sensitivity analysis without the surrogate measures of disease severity (WHO functional class and six-minute walk distance) to confirm that our findings were not driven solely by severity of disease. For longitudinal data, mixed effects generalized linear models with random intercept and slopes were fitted to account for the repeated measures of HRQoL over time. These models were adjusted for the same variables as above and interaction terms for PTE status with time and six-minute walk distance were added to account for the impact of PTE. Negative binomial regression models with an offset for time were used to determine the incidence rate ratios for hospitalization among patients with IPAH and CTEPH, adjusting for PTE status, age, sex, BMI, six-minute walk distance and WHO functional class. For this analysis, admission for PTE was not considered as a hospitalization event. We conducted a sensitivity analysis where we excluded CTEPH patients who underwent PTE from the hospitalization analysis to confirm our findings were not secondary to the PTE.

Within the CTEPH subgroup, unadjusted mixed effects generalized linear regression models were used to compare repeat measures of HRQoL between patients who had undergone PTE versus those that were medically managed. Adjusted models for repeat measures were not performed due to the overall small number of patients who underwent PTE in this cohort. All statistical analyses were performed in R version 4.0.4 and RStudio version 1.4.1106.

## Results

### Clinical Characteristics

There were 1,361 patients enrolled in PHAR by September 2019, of whom 630 met criteria for inclusion in our analysis (Figure 1). Of these, 73% had IPAH, and 27% had CTEPH. Table 1 details the baseline characteristics of the study cohort. At the time of enrollment, patients with IPAH were younger with a mean age of 55 years, more likely to be female (76%), and majority non-Hispanic white (71%). On the other hand, patients with CTEPH were older with a mean age of 58 years, 50% were female, and had a higher proportion of African Americans when compared to the IPAH group (24% vs. 11%). There were no differences in levels of education, household income, employment, and marital status between groups.

**Figure 1:**
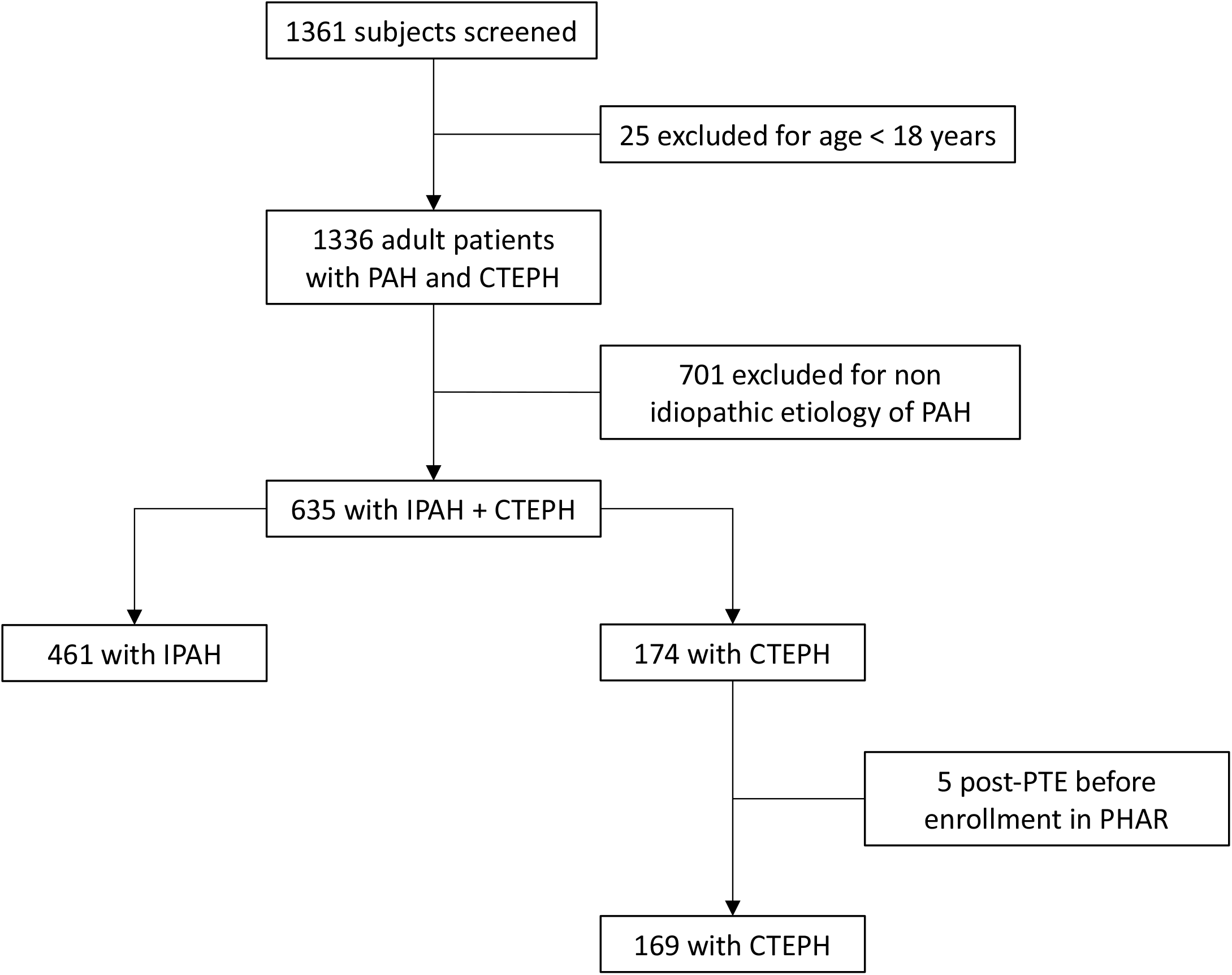
Flow diagram for study participants

**Table 1:** Patient characteristics of the cohort at time of enrollment in PHAR.

|  | IPAH<br>N = 461 | CTEPH<br>N= 169 |
| --- | --- | --- |
| Age | 55 ± 17 | 58 ± 16 |
| Female sex | 348 (76.0) | 84 (50.0) |
| BMI kg/m<sup>2</sup> (n = 614) | 31.0 ± 7.8 | 31.7 ± 8.5 |
| Race/Ethnicity |  |  |
| Non-Hispanic white | 328 (71.1) | 111 (65.7) |
| African American | 52 (11.3) | 41 (24.3) |
| Hispanic | 47 (10.2) | 9 (5.3) |
| Asian | 9 (2.0) | 0 (0.0) |
| Other/Multi-racial | 25 (5.4) | 8 (4.7) |
| Highest level of education (n=628) |  |  |
| Some Schooling | 242 (53.0) | 88 (52.7) |
| Completed high school | 172 (37.6) | 67 (40.1) |
| College or graduate studies | 43 (9.4) | 12 (7.2) |
| Household Income (n=508) |  |  |
| <\$25,000 | 100 (26.7) | 30 (23.1) |
| \$25,000 – 50,000 | 84 (22.5) | 34 (26.2) |
| \$50,000 - \$75,000 | 58 (15.5) | 20 (15.4) |
| \$75,000 – \$100,000 | 41 (11.0) | 23 (17.7) |
| >\$100,000 | 91 (24.3) | 23 (17.7) |
| Employment (n=632) |  |  |
| Homemaker / Retired | 320 (69.7) | 106 (63.1) |
| Employed (Full or part time) | 126 (27.5) | 60 (35.7) |
| Unemployed | 13 (2.8) | 2 (1.2) |
| Marital Status |  |  |
| Married / living with partner | 272 (59.0) | 95 (56.2) |
| Widowed/divorced/separated | 99 (21.5) | 41 (24.3) |
| Never married | 78 (16.9) | 30 (17.8) |
| Did not answer | 12 (2.6) | 3 (1.8) |
| Right heart catheterization hemodynamics |  |  |
| RA pressure (mm Hg) | 10 [6, 14] | 9 [7, 12] |
| Mean PA pressure (mm Hg) | 51 [42, 60] | 45 [37, 53] |
| PCWP (mm Hg) | 11 [7, 14] | 11 [8, 16] |
| Cardiac output (L/min) | 4.2 [3.2, 5.1] | 4.5 [3.6, 5.4] |
| Cardiac Index (L/min/m<sup>2</sup>) | 2.1 [1.7, 2.6] | 2.3 [1.9, 2.7] |
| PVR (Wood units) | 9.7 [6.7, 14.0] | 7.9 [5.4, 10.6] |
| Incident case | 233 (50.7) | 85 (50.9) |
| Pulmonary vasodilators |  |  |
| PDE5 inhibitors | 200 (43.4) | 7 (4.1) |
| ERA | 160 (34.7) | 8 (4.7) |
| Parenteral prostacyclin | 78 (16.9) | 1 (0.6) |
| Inhaled prostacyclin | 5 (1.1) | 0 (0.0) |
| Selexipag | 11 (2.4) | 2 (1.2) |
| Riociguat | 12 (2.6) | 37 (21.9) |
| Any combination | 176 (38.2) | 6 (3.6) |
| Baseline oxygen supplementation | 140 (30.4) | 38 (22.5) |
| Anticoagulation at presentation | 79 (17.1) | 91 (53.8) |
| Anticoagulation on follow-up | 160 (34.7) | 160 (94.7) |
| WHO functional class |  |  |
| I | 50 (11.7) | 9 (6.0) |
| II | 144 (33.8) | 66 (43.7) |
| III | 206 (48.4) | 66 (43.7) |
| IV | 26 (6.1) | 10 (6.6) |
| 6MWD (meters) | 345 ± 141 | 336 ± 118 |
| Health Related Quality of life measures |  |  |
| EmPHasis-10 | 24 ± 12 | 26 ± 12 |
| SF-12 physical component | 34 ± 7 | 33 ± 7 |
| SF-12 mental component | 49 ± 8 | 49 ± 9 |

Data are presented as N (%), mean ± standard deviation, median [Interquartile Range]. 6MWD = six-minute walk distance, RA = right atrial, PA = pulmonary artery, PCWP = pulmonary capillary wedge pressure, PVR = pulmonary vascular resistance, SF = short form, PDE5 = phosphodiesterase-5, ERA = endothelin receptor antagonists

When compared to the IPAH group, patients with CTEPH had lower median mean pulmonary artery pressures (45 vs. 51 mm Hg), lower pulmonary vascular resistance (7.9 vs. 9.7 Woods units), and higher cardiac output (4.5 vs. 4.2 L/min) but similar cardiac indices on their initial right heart catheterization. There were no differences in the WHO functional class or six-minute walk distance between the groups. Patients with CTEPH were most frequently treated with riociguat and anticoagulation therapy, whereas patients with IPAH were most often on phosphodiesterase-5 inhibitors, endothelin receptor antagonists, or a combination of both. At the time of referral to a PHAR enrolling site, only 54% of CTEPH patients were receiving anticoagulation; however, on follow-up 95% were on anticoagulation therapy.

### Health related quality of life measures

At the time of enrollment, there was no difference in unadjusted mean E10 scores between patients with CTEPH and IPAH (Table 1). However, CTEPH patients had higher E10 scores than patients with IPAH after adjusting for age, sex, BMI, six-minute walk distance and WHO functional class (β = 2.83, SE = 1.11, p = 0.01) (Figure 2). There were no differences in the baseline mental or physical components of the SF-12 among the two groups even after multivariable adjustment (Figure 2). Over time, there were no differences in either HRQoL score among the two groups, even after adjustment for PTE status (Figure 3). Sensitivity analyses excluding surrogate measures of disease severity (six-minute walk distance and WHO functional class) had similar findings. There were no differences in SF-12 scores over time between the two groups.

**Figure 2:**
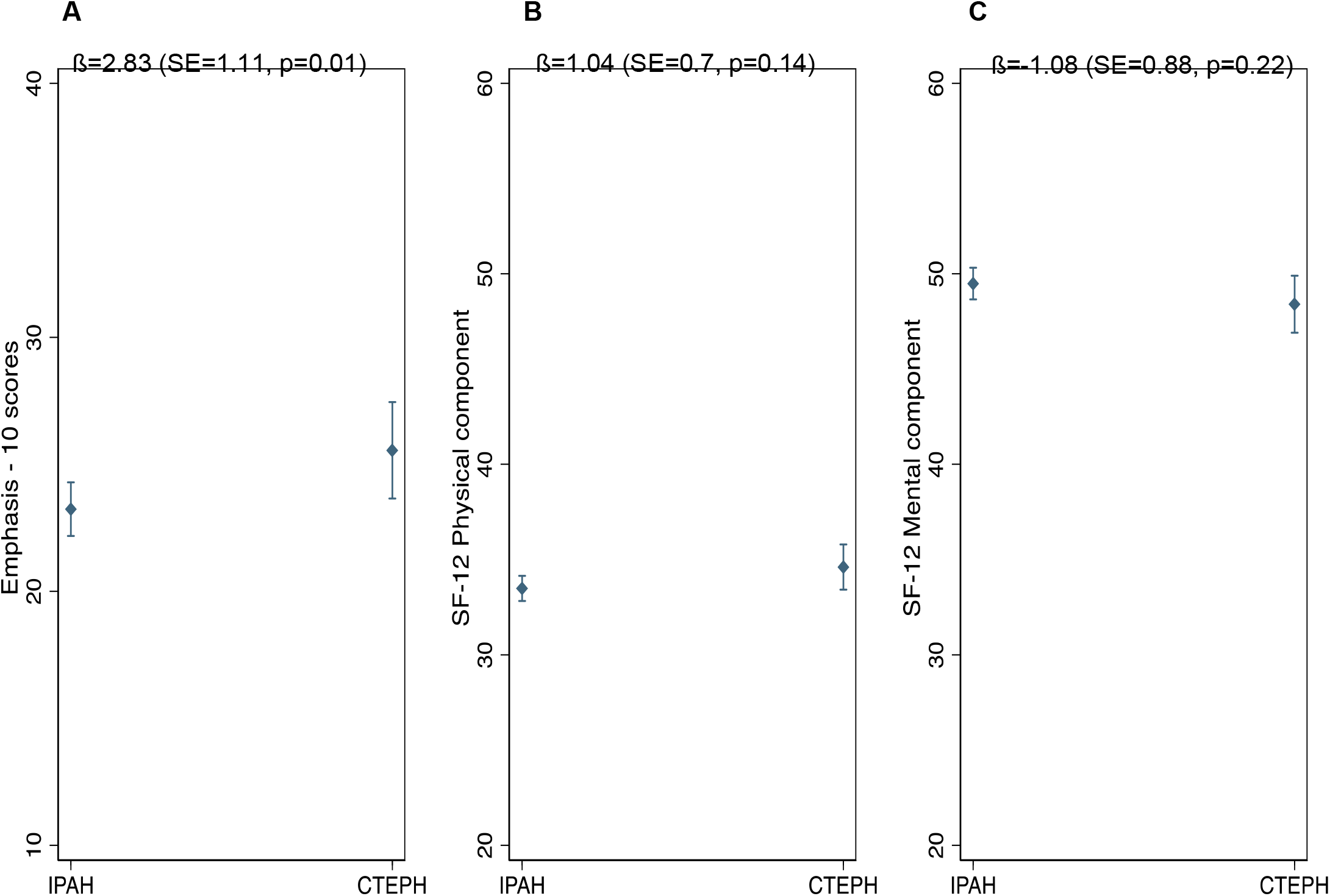
Expected mean estimates for patients with IPAH vs. CTEPH of (A) emPHasis-10 scores, (B) short form-12 physical component and (C) short form-12 mental component scores at the time of enrollment adjusted for age, sex, BMI, six-minute walk distance and WHO functional class

**Figure 3:**
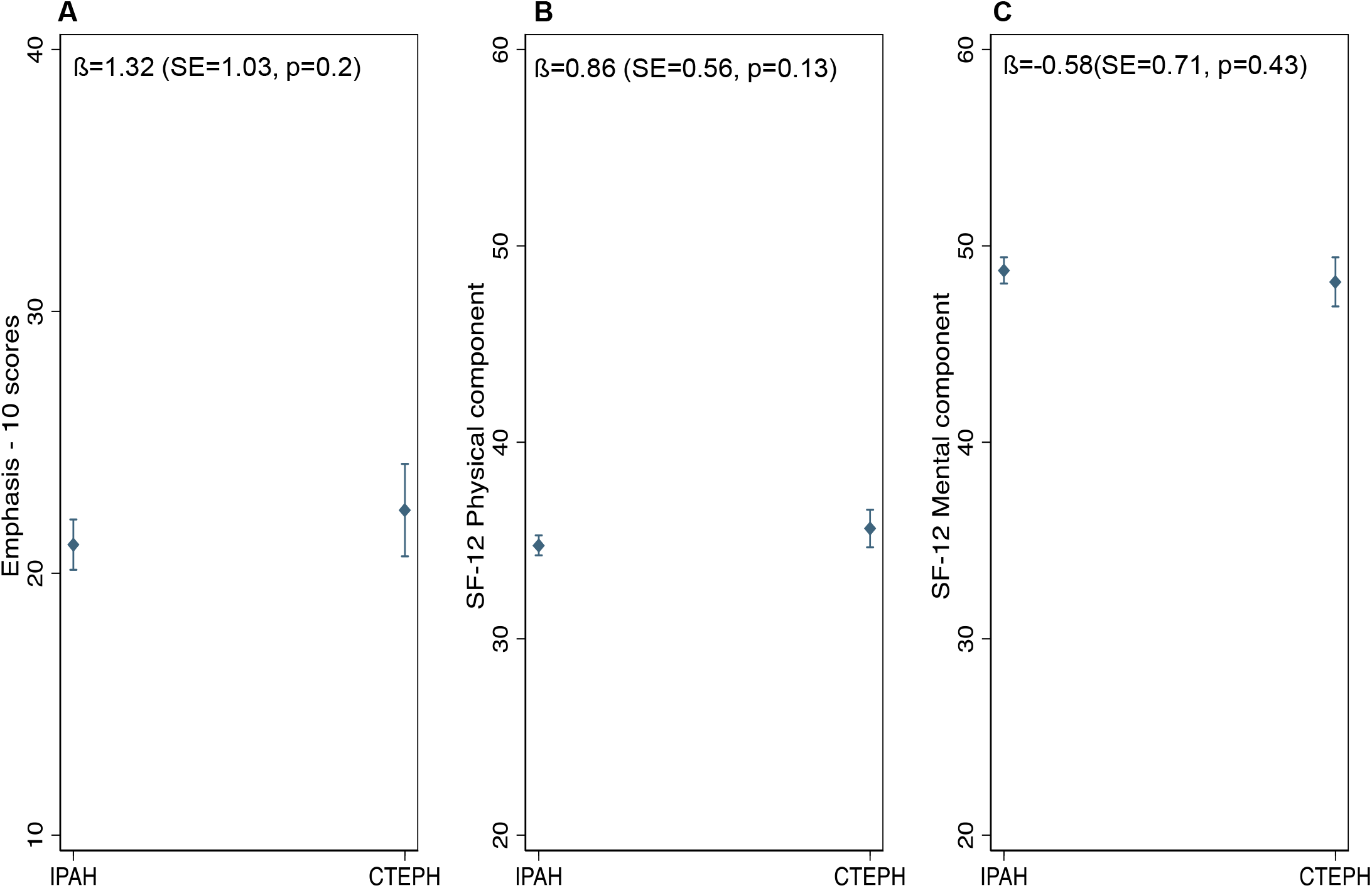
Expected mean estimates for patients with IPAH vs. CTEPH of (A) emPHasis-10 scores, (B) short form -12 physical component and (C) short form-12 mental component scores over time adjusted for age, sex, BMI, six-minute walk distance, WHO functional class, and pulmonary thromboendarterectomy status

### Hospitalizations

Of the 630 patients included in our study, 515 patients had at least one subsequent follow-up visit. During 1,585 person-years of follow-up, there were 939 hospitalizations recorded. Patients with CTEPH had a higher incidence rate ratio of all-cause hospital admissions than patients with IPAH (IRR 1.66, 95%CI 1.04 – 2.65, p = 0.035) after adjustment for age, sex, BMI, six-minute walk distance, WHO functional class and PTE status (Figure 4). In sensitivity analysis excluding CTEPH patients who underwent PTE, CTEPH patients were again noted to have increased incidence rates of hospitalizations as compared to IPAH patients (IRR 1.90, 95%CI 1.08-3.40, p = 0.027).

**Figure 4:**
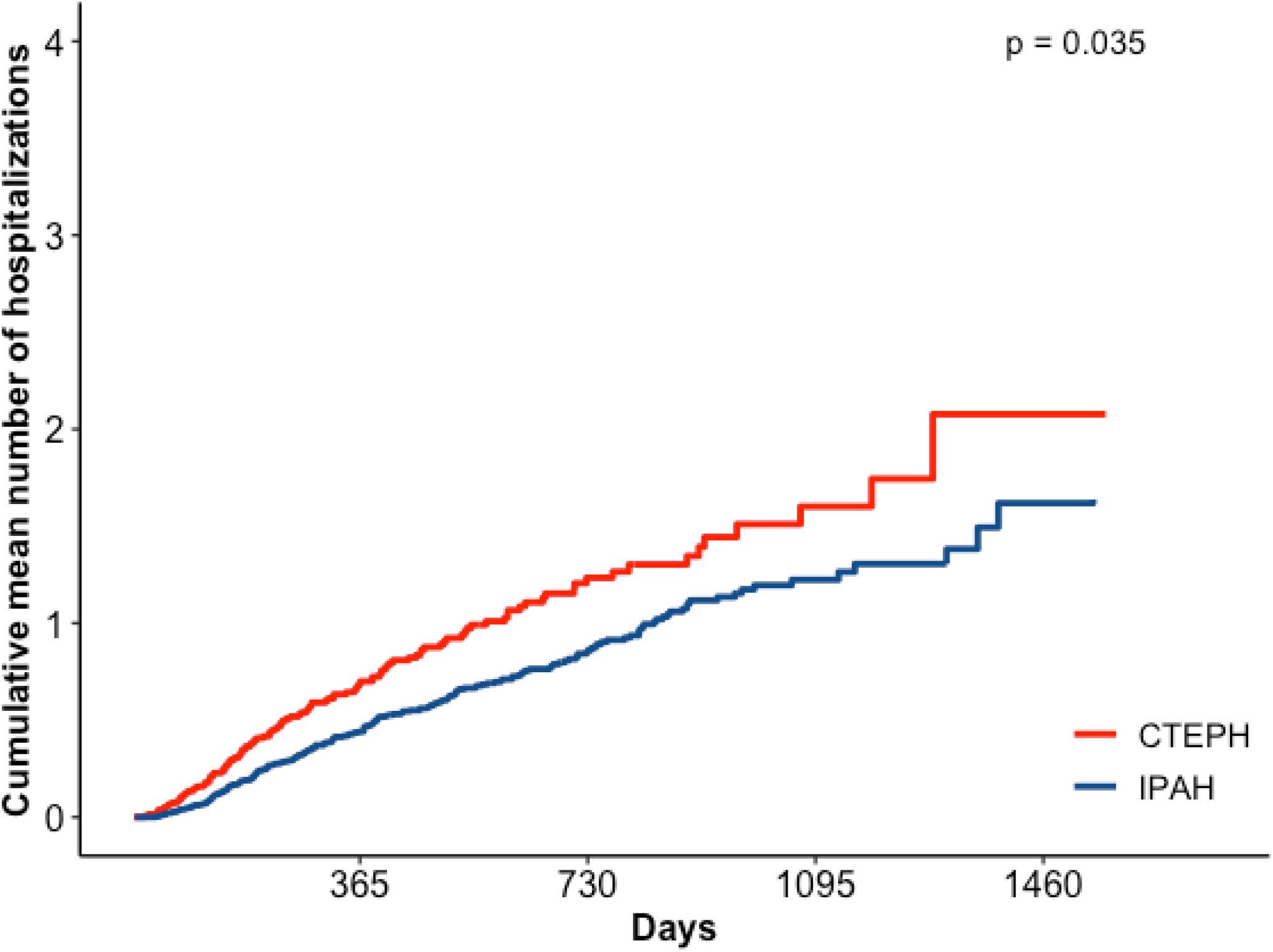
Cumulative mean hospitalizations over time in patients with chronic thromboembolic pulmonary hypertension (CTEPH) versus those with idiopathic pulmonary arterial hypertension (IPAH)

### CTEPH subgroup analysis

In PHAR, 63% of patients with CTEPH were referred for surgical evaluation at enrollment. By the end of follow up 22% had undergone PTE and 78% were managed medically. Mean age was 59 years and majority females (69%) among all patients who were medically managed throughout and did not undergo PTE (Table 2). The CTEPH patients who underwent PTE were younger with a mean age of 54 years and more likely to be male (62%). There were no differences in levels of education, household income, employment, or marital status between the two groups (Table 2).

**Table 2:** Baseline characteristics of CTEPH patients who were medically managed or subsequently underwent pulmonary thromboendarterectomy (PTE)

|  | Medical<br>management<br>N = 133 | PTE group<br>N = 36 |
| --- | --- | --- |
| Age | 59 ± 16 | 54 ± 15 |
| Female sex | 69 (52.0) | 64 (38.0) |
| BMI kg/m<sup>2</sup> | 31.3 ± 8.0 | 30 ± 7.7 |
| Race/Ethnicity |  |  |
| Non-Hispanic white | 85 (64) | 26 (72) |
| African American | 34 (25.6) | 7 (19.4) |
| Hispanic | 8 (6.0) | 1 (2.8) |
| Asian | -- | -- |
| Other/Multi-racial | 4 (4.5) | 2 (5.6) |
| Highest level of education |  |  |
| Some Schooling | 70 (53) | 18 (51) |
| Completed high school | 52 (39) | 15 (43) |
| College or graduate studies | 10 (8) | 2 (6) |
| Household Income |  |  |
| <\$25,000 | 24 (24) | 6 (21) |
| \$25,000 – 50,000 | 25 (25.7) | 8 (28) |
| \$50,000 - \$75,000 | 12 (11.9) | 8 (27.6) |
| \$75,000 – \$100,000 | 21 (20.8) | 2 (6.9) |
| >\$100,000 | 18 (17.8) | 5 (17.2) |
| Employment |  |  |
| Homemaker / Retired | 85 (64.4) | 21 (58.3) |
| Employed (Full or part time) | 45 (34.1) | 15 (41.7) |
| Unemployed | 2 (1.5) | -- |
| Marital Status |  |  |
| Married / living with partner | 77 (57.9) | 18 (50) |
| Widowed/divorced/separated | 32 (24.1) | 9 (25) |
| Never married | 21 (15.8) | 9 (25) |
| Did not answer | 3 (2.3) | -- |
| Right heart catheterization hemodynamics |  |  |
| RA pressure (mm Hg) | 9 [6, 13] | 10 [7, 12] |
| Mean PA pressure (mm Hg) | 45 [37, 54] | 45 [42, 51] |
| PCWP (mm Hg) | 11 [8, 16] | 11 [7, 14] |
| Cardiac output (L/min) | 4.3 [3.52, 5.4] | 4.7 [4.2, 5.5] |
| Cardiac Index (L/min/m<sup>2</sup>) | 2.22 [1.88, 2.56] | 2.29 [1.90, 2.80] |
| PVR (Wood units) | 8 [5, 11] | 7 [5, 10] |
| Pulmonary vasodilators |  |  |
| PDE5 inhibitors | 7 (5.3) | -- |
| ERA | 8 (6.0) | -- |
| Parenteral prostacyclin | 1 (0.8) | -- |
| Inhaled prostacyclin | -- | -- |
| Selexipag | 2 (1.5) | -- |
| Riociguat | 31 (23.3) | 6 (16.7) |
| Any combination | 6 (4.5) | -- |
| Baseline oxygen supplementation | 29 (21.8) | 9 (25.0) |
| Anticoagulation at presentation | 61 (45.9) | 30 (83.3) |
| Anticoagulation on follow-up | 125 (94.0) | 35 (97.2) |
| WHO functional class |  |  |
| I | 9 (7.6) | -- |
| II | 55 (46.6) | 11 (33.3) |
| III | 45 (38.1) | 21 (63.6) |
| IV | 9 (7.6) | 1 (3.0) |
| 6MWD (meters) | 335 ± 138 | 425 ± 82 |
| Health Related Quality of life measures |  |  |
| EmPHasis-10 | 24 ± 12 | 29 ± 11 |
| SF-12 physical component | 34 ± 7 | 35 ± 6 |
| SF-12 mental component | 49 ± 8 | 47 ± 9 |

Data are presented as N (%), mean ± standard deviation, median [Interquartile Range]. 6MWD = six-minute walk distance, RA = right atrial, PA = pulmonary artery, PCWP = pulmonary capillary wedge pressure, PVR = pulmonary vascular resistance, SF = short form, PDE5 = phosphodiesterase-5, ERA = endothelin receptor antagonists

When compared to those that were medically managed, patients who underwent PTE had a higher cardiac output (4.7 vs. 4.3 L/min) and higher E10 scores (worse HRQoL, 29 vs. 24) but were less likely to present in WHO functional class IV (3% vs. 6%) and had greater six-minute walk distances (425 vs. 335 meters). There were no differences in cardiac indices, right atrial pressure, pulmonary artery pressure, pulmonary capillary wedge pressure between the groups and both were most frequently treated with riociguat (Table 2).

Patients who underwent PTE had a decrease in their E10 scores postoperatively indicating improved HRQoL (Figure 5). Unadjusted mixed effects models showed that patients who underwent PTE had lower E10 scores and higher SF-12 physical component scores, both indicative of better HRQoL (Figure 6). There were no differences in SF-12 mental component scores between the two subgroups.

**Figure 5:**
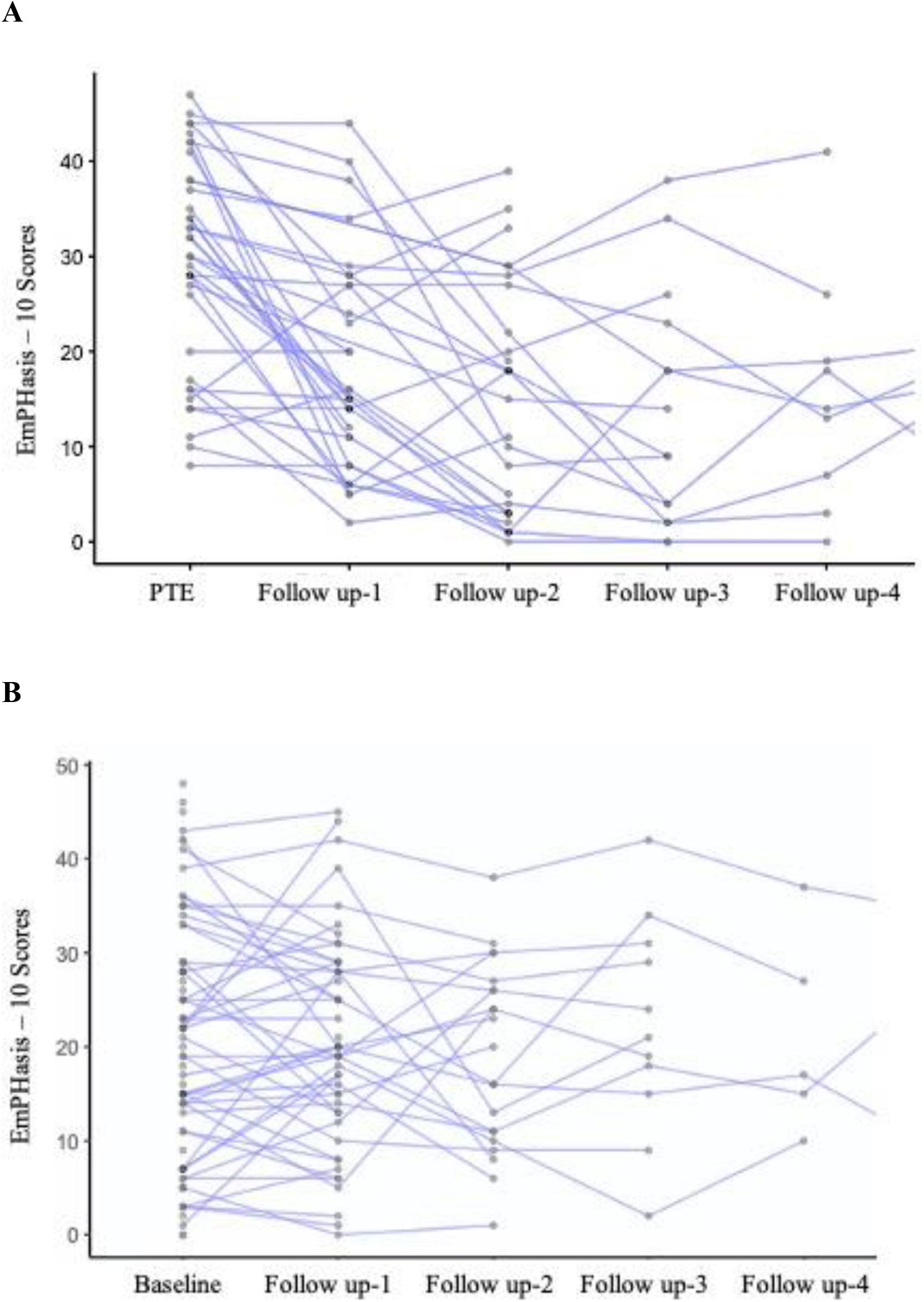
Spaghetti plot of EmPHasis-10 scores for individual patients with chronic thromboembolic pulmonary hypertension (CTEPH) (A) post-pulmonary thromendarterectomy (PTE) (B) medical management only

**Figure 6:**
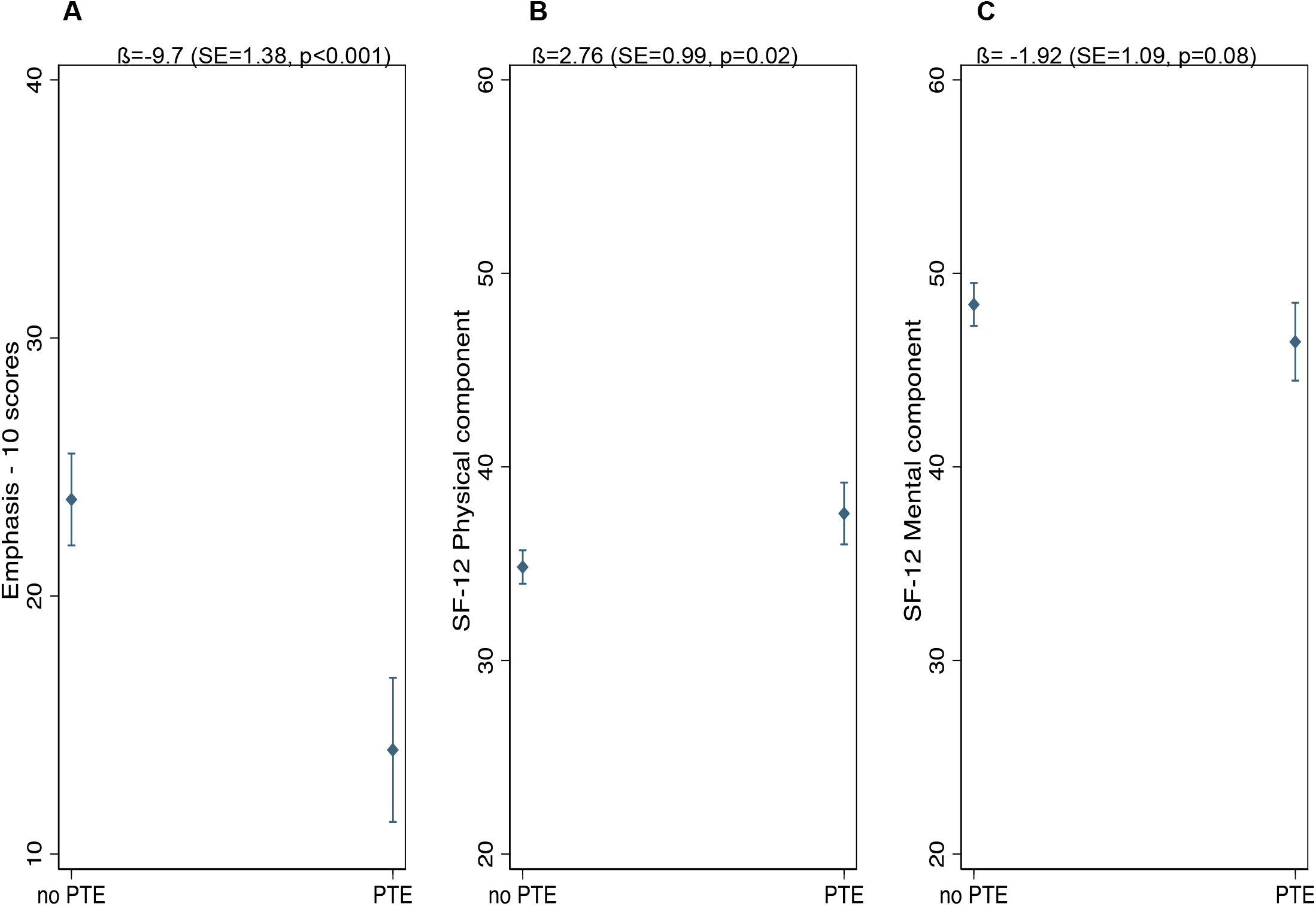
Expected unadjusted mean estimates for patients with CTEPH who underwent pulmonary pulmonary thromendarterectomy (PTE) vs. those who were medically managed of (A) emPHasis-10 scores, (B) short form -12 physical component and (C) short form-12 mental component scores over time.

## Discussion

In a large multicenter prospective cohort of patients with IPAH and CTEPH across the U.S., patients with CTEPH at enrollment were older, with better hemodynamics (lower pulmonary artery systolic pressure and pulmonary vascular resistance) but worse HRQoL when compared to patients with IPAH after adjustment for baseline variables; however, the difference in HRQoL did not persist over time. Patients with CTEPH had higher hospitalization rates (after excluding hospitalization for PTE) than patients with IPAH. Among CTEPH patients, those who eventually underwent PTE were younger, male, with a higher cardiac output and better six-minute walk distance than those who were medically managed. In addition, patients with CTEPH who underwent PTE had improvement in their HRQoL.

Pulmonary hypertension is a rare debilitating disease that negatively impacts the quality of life of patients and their caregivers. In addition to hemodynamic measurements, laboratory biomarkers and exercise testing, there has been increased recognition of the importance of incorporating patient-centered outcomes into management decisions and clinical trial design.(11, 19, 20) A patient-reported outcome is a self-assessment of health by patients and frequently includes HRQoL.(21, 22) Several studies have demonstrated a strong correlation between patient reported HRQoL and outcomes such as the six-minute walk distance, hospitalizations and survival in pulmonary hypertension.(3, 23, 24) Studies exploring differences in HRQoL between patients with CTEPH and IPAH have mixed results with some finding no difference and others showing CTEPH patients having worse HRQoL.(13, 25, 26) The results from this study demonstrate that patients with CTEPH have worse HRQoL at referral to a Pulmonary Hypertension Care Center as compared to patients with IPAH, but these differences do not persist over time in a large prospective multicenter cohort. Among CTEPH patients, similar to what has been previously reported,(26) this study found that patients who undergo PTE have significant improvement in their HRQoL, and the improvement in scores may be more significant in patients without residual disease post-PTE.(4, 5)

The worse HRQoL observed in CTEPH patients at referral was not explained by older age as compared to IPAH patients; however, older age may be associated with increased co-morbid conditions, which were not available for us to account for. Over time, with treatment, both groups had similar HRQoL scores.

The improvement in the CTEPH group may be driven by improved HRQoL scores in the subgroup of CTEPH patients who underwent PTE. In contrast, medically managed patients with CTEPH and IPAH patients continued to have similar HRQoL. Since most patients with CTEPH in this cohort were medically managed, this may have biased our results towards the null. While this study found differences in E-10 scores, no differences were detected in SF-12 scores among patients with IPAH versus those with CTEPH. The E-10 is a PH-specific instrument and may thus be a more sensitive tool to measure HRQoL in these patient populations.

PTE is a potentially curative procedure in CTEPH and is recommended as the treatment of choice in this population by the European Society of Cardiology/European Respiratory Society expert consensus guidelines.(27, 28) Successful PTE has been associated with lower right atrial pressure, higher cardiac index, better WHO functional class, and improved survival.(4, 29) In the overall population of patients with CTEPH, it is estimated that PTE is feasible in 63 – 76% of patients and 71 – 88% of those who are deemed operable undergo the procedure.(30) At their initial assessment at a Pulmonary Hypertension Care Center only 63% of patients with CTEPH were referred for surgical evaluation for PTE. The relatively low rate of referral to PTE at enrollment in the PHAR is likely a reflection of their incident case status and referrals often occur after evaluation at one of the Pulmonary Hypertension Care Centers. Of all the CTEPH pateitns in our cohort, 78% were medically managed, and only 22% underwent PTE. An additional 5 patients had undergone PTE prior to enrollment in the PHAR and were excluded from our analysis. In our cohort, rates of PTE were significantly lower presumably because patients either declined surgery or had factors that deemed them unsuitable for surgery – such as co-morbidities or distribution of clots on imaging. Reasons for no surgical referral and reasons for no PTE after surgical referral were not recorded in the PHAR. Due to the low number of patients who underwent PTE in this cohort, we were unable to examine how hospitalization rates change post PTE.

Pulmonary hypertension is associated with higher health care utilization and costs.(9, 10, 31, 32) Studies of patients with CTEPH found that these patients have up to 6-times higher inpatient and outpatient costs and overall health care utilization than matched controls.(8, 31) However, we are not aware of prior studies comparing hospitalization rates between IPAH and CTEPH. In our cohort, patients with CTEPH had higher hospitalization rates than those with IPAH (excluding hospitalization for PTE) even after adjusting for patient demographics and disease severity. It is possible that this may again be a reflection of the higher rate of associated co-morbidities in this patient cohort, or may include admissions for balloon pulmonary angioplasty, data for which were not collected in PHAR at the time of analysis.

This study has some limitations. Seven percent of patients in this registry were lost to follow up. Based on the “real world” registry protocol, patients were seen in follow up “as clinically needed” in approximately six-month intervals, thus resulting in variable follow up times for patients. However, this should not lead to a bias due to the non-differential nature of the missing data. Additionally, all of the previously presented longitudinal analyses incorporated patient follow-up times. Details of comorbid conditions and pulmonary vascular imaging were not recorded in the registry, limiting the assessment of surgical eligibility. Additionally, data on balloon pulmonary angioplasty was not collected in this cohort during the study period. Finally, the overall rates of PTE were lower in this cohort than those generally reported in the CTEPH population, but data on lack of surgical eligibility, patient refusal and data on balloon pulmonary angioplasty were not captured. To address these limitations, the PHAR began prospectively recording data on balloon pulmonary angioplasties for patients starting in June 2020.

Future studies will focus on the inclusion of this data for analyses. In a large, multicenter, prospective cohort, patients with CTEPH were found to have significantly worse HRQoL at initial presentation to pulmonary hypertension specialty centers compared to patients with IPAH; however, these differences did not persist over time. CTEPH patients who underwent PTE experienced significant improvements in HRQoL. The worse baseline quality of life and higher hospitalization rate of CTEPH patients presents an opportunity for improvement in clinical management of these patients.

## Data Availability

The data that support the findings of this study are available from the pulmonary hypertension association registry, but restrictions may apply. The datasets generated during and/or analyzed during the current study are available from the corresponding author upon reasonable request and with permission from the Pulmonary Hypertension Association Registry.

## Acknowledgements

The Pulmonary Hypertension Association Registry (PHAR) is supported by Pulmonary Hypertension Care Centers, Inc., a supporting organization of the Pulmonary Hypertension Association. The authors thank the other investigators, the staff, and particularly participants of the PHAR for their valuable contributions. A full list of participating PHAR sites and institutions can be found at www.PHAssociation.org/PHAR.”

## Appendix – PHAR Investigators

Abhijit Raval, MD, Amresh Raina, MD, Anna Hemnes, MD, Bruce Andrus, MD, Charles Burger, MD, Corey Ventetuolo, MD, D. Dunbar Ivy, MD, Delphine Yung, MD, Eric Austin, MD, Eric Roberts, MD, Erika Berman-Rosenzweig, MD, Gautam Ramani, MD, Granthem Farr, MD, H. James Ford, MD, James Klinger, MD, James Runo, MD, Jeff Fineman, MD, Jessica Huston, MD, John Swisher,MD, PhD, Kenneth Presberg, MD, Kishan Parikh, MD, Lana Melendres-Groves, MD, Linda Cadaret, MD, Mark Avdalovic, MD, Michael Duncan, MD, Murali Chakinala, MD, Nidhy Varghese, MD, Paul Boyce, MD, Peter Leary, MD, PhD, R. James White, MD, PhD, Rahul Argula, MD, Rana Awdish, MD, Raymond Foley, DO, Roham Zamanian, MD, Russel Hirsch, MD, Sahil Bakshi, MD, Sapna Desai, MD, Steven Kawut, MD, MS, Tammy Wichman, MD, Timothy Williamson, MD.

